# Acute Ischemic Stroke Detection on Non-Contrast CT: A Deep Learning Approach

**DOI:** 10.64898/2026.06.20.26356152

**Authors:** Ansh Goyal, Robert David Stevens

**Affiliations:** Laboratory of Computational Intensive Care Medicine, Johns Hopkins University School of Medicine, Baltimore, Maryland, United States of America; Department of Biomedical Engineering, Whiting School of Engineering, Johns Hopkins University, Baltimore, Maryland, United States of America; Department of Electrical and Computer Engineering, Whiting School of Engineering, Johns Hopkins University, Baltimore, Maryland, United States of America; Department of Anesthesiology and Critical Care Medicine, Johns Hopkins University School of Medicine, Baltimore, Maryland, United States of America; Department of Radiology and Radiological Science, Johns Hopkins University School of Medicine, Baltimore, Maryland, United States of America

**Keywords:** Acute ischemic stroke, non-contrast CT, deep learning, classification, segmentation, small lesions

## Abstract

Acute ischemic stroke (AIS) is a leading cause of disability and death while effective treatment requires quick and accurate diagnosis. Non-contrast CT (NCCT) is widely used in the initial screening of AIS, but stroke detection is challenging because early changes on NCCT are subtle or indistinguishable. Using hyperacute NCCTs as inputs and diffusion-weighted MRI as ground truth, we trained a deep learning algorithm to classify patients with AIS and segment the stroke lesions. We hypothesized that this approach would accurately detect hyperacute tissue density changes on NCCT. For the classification task, our ResNet50 model delivered the best performance (with 98.5% accuracy, 97.4% precision, and 100% recall on an evaluation set). Classification performance remained strong when restricted to lesions smaller than 5 mL, which constituted the majority of our evaluation cases. For the segmentation task accomplished using a range of U-Net architectures, performance was acceptable for large lesions and declined sharply for smaller lesions. Together, these findings demonstrate the feasibility of deep learning for AIS detection and represent a step towards faster triage and treatment for stroke patients.

## 1. Introduction

Acute ischemic stroke is a leading cause of long-term disability and death worldwide. In the acute phase, treatments like intravenous thrombolysis and mechanical thrombectomy are highly effective when delivered within a narrow temporal window following symptom onset. Non-contrast computed tomography (NCCT) is widely used in the initial evaluation of patients with AIS-like presentations as it allows rapid and accurate exclusion of other etiologies such as intracerebral hemorrhage. However, the early identification of ischemic lesions is challenging on NCCT because brain parenchymal changes may be subtle or indistinguishable, in particular when the infarct volume is small [1]. In current workflows NCCT is generally supplemented with CT angiography (CTA) and CT perfusion (CTP) imaging or diffusion-weighted brain MRI (DWI) which have higher specificity for stroke detection and can guide treatment decisions, however these techniques impose additional delays and may not be available in lower-resource settings [2]. There is consequentially significant interest in alternative approaches which would allow rapid AIS detection and localization.

Since NCCT is the first-line imaging modality in AIS evaluation, a stroke detection algorithm leveraging NCCT could be highly impactful. Recent studies have demonstrated the potential of deep learning for detection and segmentation of various stroke types on NCCT [3, 4, 5, 6]. While these works represent meaningful advances in NCCT-based stroke analysis, they primarily focus on larger, more visible lesions and do not directly address model performance in the small-volume, early-stage infarcts. A clinically viable model must demonstrate robust performance on small lesions and low contrast, a challenge that remains largely unaddressed.

Two primary tasks for image based AIS evaluation are classification and segmentation. Classification aims to identify the presence of a stroke in the given scan, while segmentation seeks to predict the exact location of the stroke. While lesion segmentation is an important objective, it is a complex challenge due to lesion variability, low contrast, and imaging noise. The ability to determine the existence of a stroke without expert intervention represents a substantial advance over current workflows.

The principal objective in this study is to determine if deep learning models trained with NCCT inputs and DWI as ground truth labels can accurately discriminate stroke from non-stroke patients. We hypothesize that this approach detects early stroke-associated tissue changes on NCCT. The secondary task is to determine if such algorithms can localize stroke lesions on NCCT. Our findings point to the feasibility of deep learning in AIS detection and lay the foundation for future advancements in automated stroke assessment.

## 2. Materials and Methods

### 2.1. Dataset

The data were curated from a prospective registry created and maintained at the Johns Hopkins Hospital (JHH) Comprehensive Stroke Center (stroke patients, N=578) and from an age- and gender- matched group of JHH inpatients (non-stroke controls, N=463). All patients were adults who underwent NCCT and MRI-DWI. All scans were acquired at JHH under a standard clinical NCCT protocol. Patient demographics and clinical characteristics for both cohorts are summarized in Table 1.

This stroke dataset is characterized by a high prevalence of small ischemic lesions. Across all labeled stroke-positive cases, lesion volume ranged from 0.007 mL to 251.5 mL, with a median volume of 2.0 mL. One case with a lesion volume of 0.007 mL was identified as a likely labeling artifact; excluding it, the remaining 577 stroke-positive cases spanned a volume range of 0.106 mL to 251.5 mL. Table 2 shows the distribution of scans by lesion size for our dataset in detail. Note that 64.9% of all stroke-positive scans had lesion volume under 5 mL.

All NCCT scans underwent a standardized preprocessing pipeline to ensure consistency across samples. First, 3D-Slicer [7] was used to remove all non-brain tissues (particularly the skull). Next, images were resampled to an isotropic resolution of 1 mm³ and registered to the MNI-152 atlas [8] for spatial alignment. CT windowing was then applied to confine Hounsfield unit values between 0 and 80, followed by pixel-wise normalization to the range [0, 1].

While the classification task required only binary stroke/non-stroke labels, the segmentation task demanded voxel-wise label maps to localize the ischemic lesions. To achieve this, lesion labels were generated using a semi-automatic DWI coregistration approach, leveraging DWI’s higher sensitivity to ischemic changes. For MRI preprocessing, we followed the procedures outlined in [9], which included resampling, skull-stripping, normalization, and alignment to standard space. From each MRI volume, key sequences (B0, ADC, and DWI) were extracted for lesion detection. The preliminary infarct maps were generated using DAGMNet [9], achieving a mean Dice coefficient above 0.75. To mitigate possible errors, neuroradiologists reviewed and refined these lesion masks for accuracy.

MRI-derived infarct labels were then coregistered with their corresponding NCCT scans using itk-Elastix [10]. Transformation matrices were applied to map lesion masks into NCCT space, accounting for translation, rotation, scaling, and shearing. The resulting co-registered NCCT–label pairs formed the basis for segmentation model training; Figure 1 summarizes the full dataset creation pipeline. For classification, the 1,041 NCCT scans were divided into training (80%, 832 scans), and evaluation (20%, 209 scans), ensuring balanced representation of stroke and non-stroke cases. For segmentation, the 578 labeled NCCT scans were split into 360 training, 102 validation, and 116 testing cases. All splits were done at a patient level to ensure no patient was seen in both training and evaluation sets.

**Figure 1.**
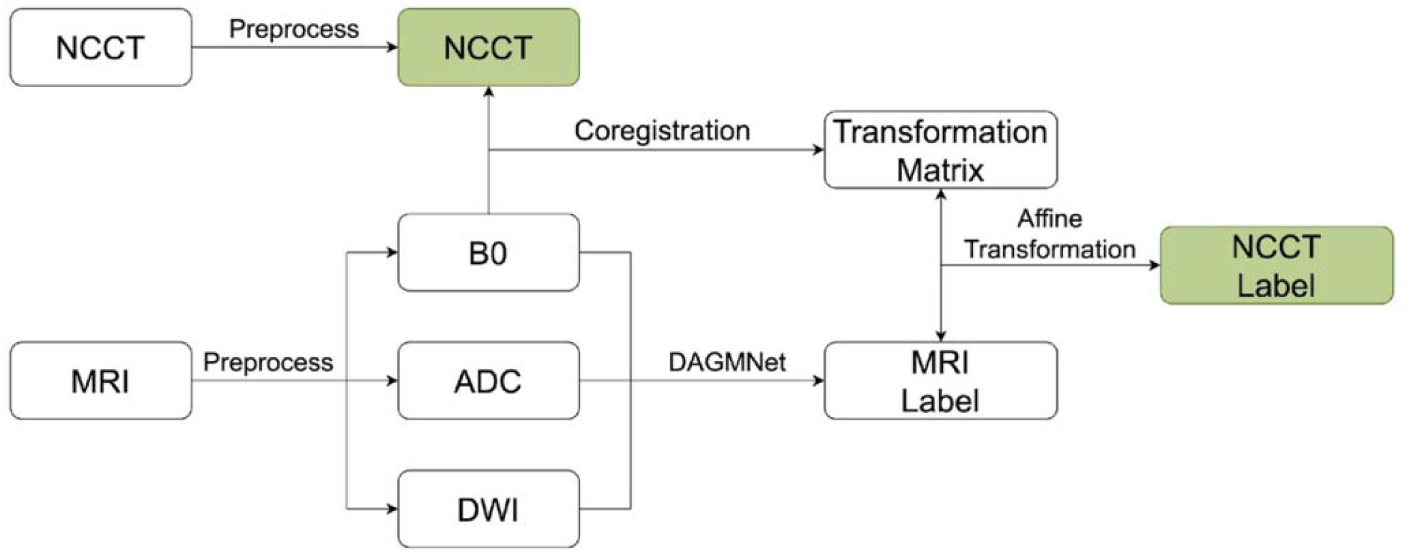
Dataset creation flowchart. Green block indicates the final co-registered NCCT–label pair used for segmentation model training.

### 2.2. Classification Models

For the classification task, we implemented two deep learning architectures:

1. **ResNet50 [11]:** ResNet50 is a 50-layer convolutional neural network that employs residual connections to mitigate vanishing gradients and facilitate the training of deeper networks. The architecture consists of bottleneck residual blocks (1×1, 3×3, 1×1 convolutions) that enable hierarchical feature extraction at high computational efficiency. The model follows a four-stage design, with global average pooling followed by a final softmax classifier for binary prediction (Figure 2).
2. **Vision Transformer (ViT) [12]:** The Vision Transformer is a self-attention–based model that treats each image as a sequence of fixed-size patches. Each patch is linearly embedded and positionally encoded before being processed by a transformer encoder composed of multi-head self-attention and feed-forward layers. A classification token aggregates the global image representation, and the output passes through a softmax layer for final prediction (Figure 3).

**Figure 2.**
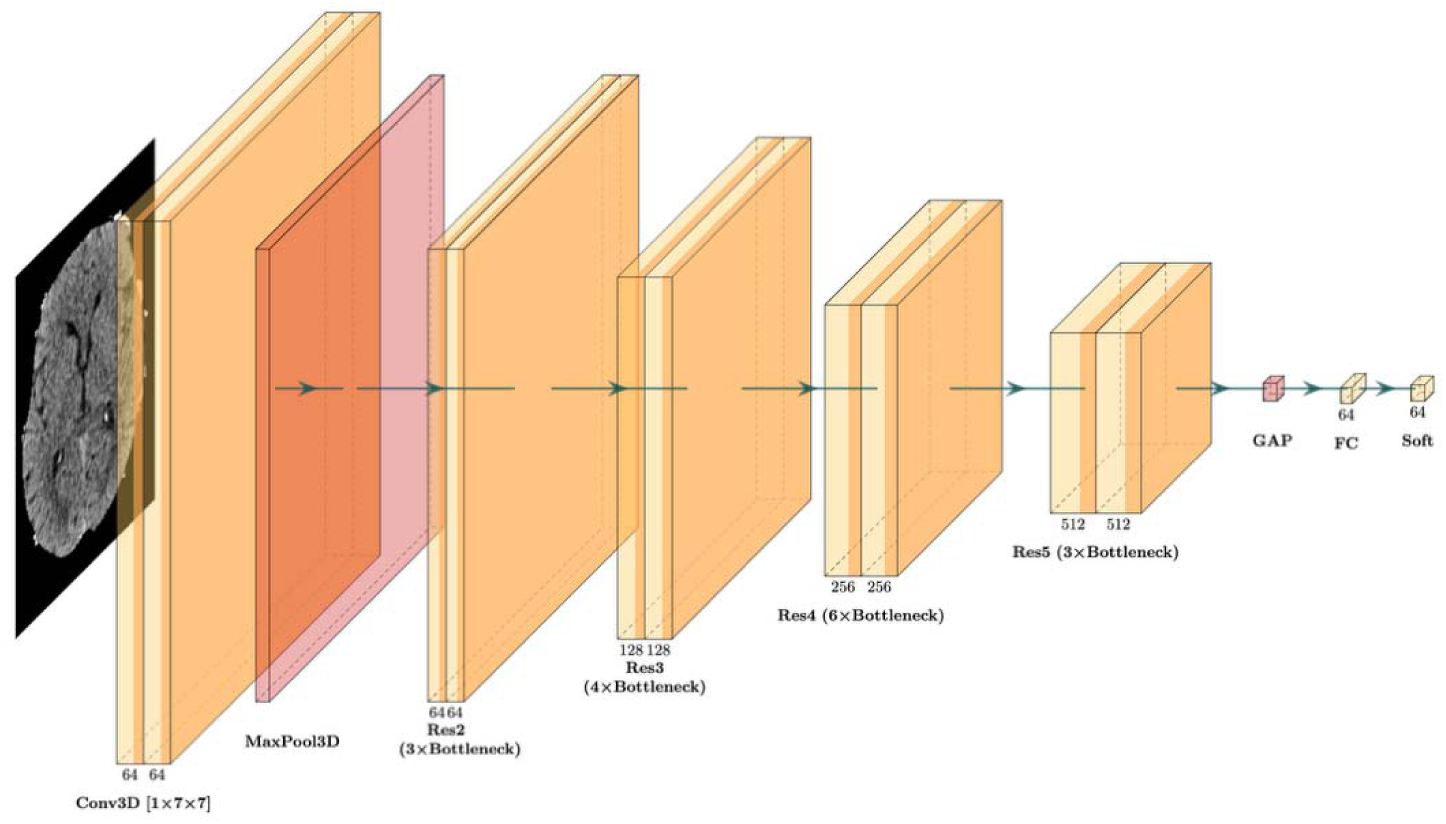
ResNet50 model architecture used for the classification task.

**Figure 3.**
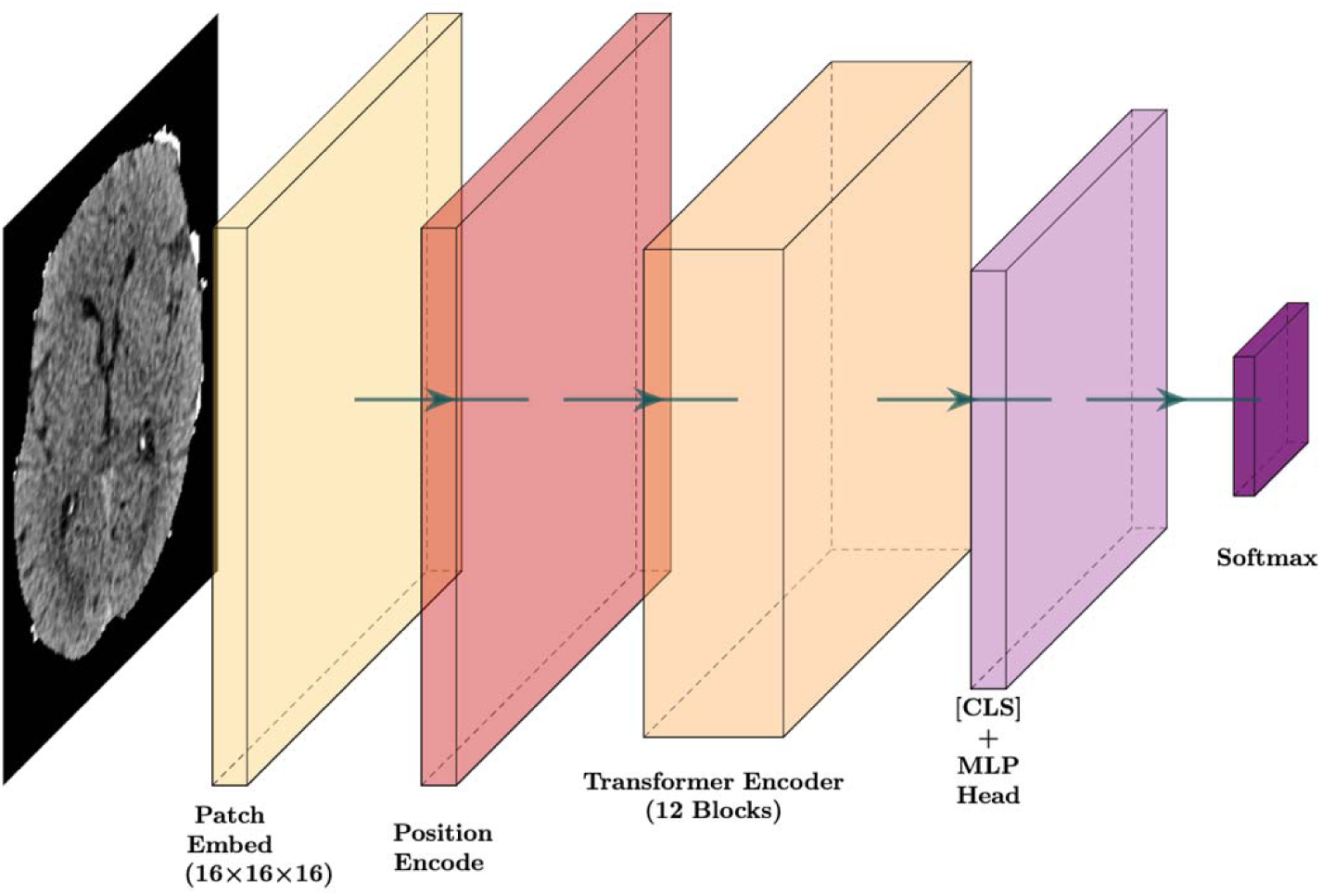
Vision Transformer (ViT) model architecture used for the classification task.

Both of these models were trained on the same dataset (as described above), and both output a binary classification (stroke-positive vs. stroke-negative).

### 2.3. Segmentation Models

To evaluate ischemic lesion segmentation on NCCT, we compared four commonly used segmentation architectures: a 3D U-Net, a dynamic U-Net, a classic attention U-Net, and our own modified attention-based U-Net (AUIS). All models were evaluated with a consistent training and evaluation framework for fair comparison.

The classic Attention U-Net [13] serves as a baseline model for medical image segmentation. Its encoder-decoder structure with skip connections enables effective feature aggregation, and it has demonstrated robust performance in many imaging modalities. The 3D U-Net [14] extends the architecture by operating fully in three dimensions, allowing for volumetric context to be utilized. The dynamic U-Net [15] adapts convolutional kernel configurations across layers, and was included as a more flexible variant to improve feature representation. All of these models were implemented in their standard architectures using the Medical Open Network for AI (MONAI) framework [16]. Details on their implementation can be found in MONAI documentation.

In addition to these standard architectures, we implemented a modified attention-based U-Net model called AUIS, inspired by the original Attention U-Net [13]. AUIS follows an encoder-bottleneck-decoder structure and has an attention mechanism gate (AMG) to enhance focus on ischemic stroke regions. The encoder processes NCCT images through input and down-sampling layers to extract high-level features. The bottleneck connects the encoder and decoders, capturing the abstract feature representations of stroke lesions. Finally, the decoder upsamples the extracted features to restore spatial resolution. To reduce memory usage and optimize for the size of our dataset, we removed one encoder and one decoder block.

The attention mechanism gate combines two input features (referred to as x1 and p1), and processes them through ReLU and Sigmoid activation functions. After activation, the features are reshaped into a heatmap that selectively enhances ischemic stroke regions. This helps focus the learning on the relevant features, minimizing background noise.

The architectural diagram provided in Figure 4 (generated using PlotNeuralNet [17]) is only for the proposed AUIS model, as the other architectures follow standard designs and were implemented directly using the publicly available MONAI implementation. By evaluating both standard and modified architectures, this study seeks to characterize the practical strengths and limitations of current approaches for NCCT-based ischemic lesion segmentation.

**Figure 4.**
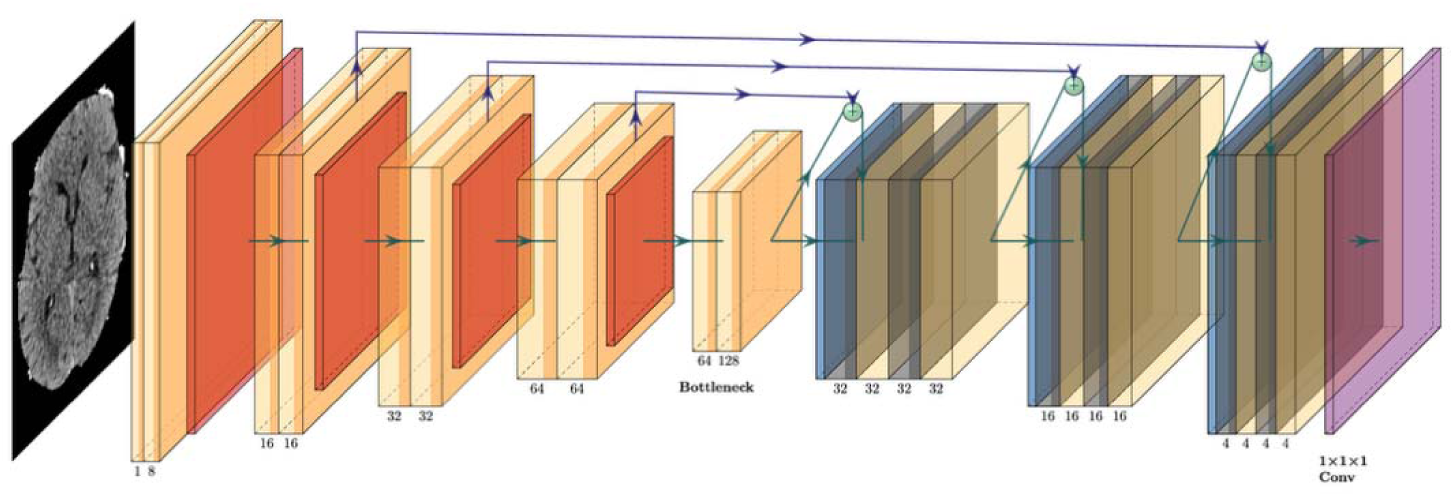
AUIS model architecture. Dark orange blocks indicate max pooling (2×2), orange blocks indicate 3×3×3 convolutions with ReLU activation, and green spheres indicate copy-and-concatenate attention gates.

### 2.4. Implementation Details

For classification, each NCCT volume was processed slice-by-slice, with each axial slice resized to 224×224 pixels and fed independently through the model. Both ResNet50 and ViT were initialized with ImageNet pretrained weights, with grayscale slices converted to three-channel inputs accordingly. Scan-level predictions were obtained by aggregating slice-level logits using log-mean-exp pooling across all slices in the volume. No data augmentation was applied. Models were trained using cross-entropy loss with a batch size of 8 for ResNet50 and 4 for ViT. For segmentation, training employed a hybrid Dice + focal loss to address class imbalance. Both tasks used the Adam optimizer with a learning rate of 1×10 . Training was conducted for 250 epochs on two NVIDIA A100 PCIe 40 GB GPUs over approximately 48 hours, and the final checkpoint was used for evaluation. For the classification task, 95% confidence intervals for the reported metrics were estimated by nonparametric bootstrap resampling of the held-out evaluation set with replacement (2,000 resamples).

During segmentation, feature maps were downsampled by a factor of (2,2,2) in each encoder block and upsampled symmetrically in the decoder. The final output was a voxel-wise probability map representing the likelihood of lesion presence. These probabilities were thresholded to generate binary masks, with the threshold set as the mean of the minimum and maximum probabilities (typically near 0.5).

The source code for the models is available on GitHub under the following link: https://github.com/AnshGoyal1123/Acute-Stroke-Detection.

## 3. Results

Table 1 shows the clinical demographic information and sample characteristics for the population evaluated in the study. For the non-stroke cohort, the most common NCCT indications were trauma-related (52.8%), routine non-contrast protocols (33.4%), and carotid CT angiography (10.3%). The specific clinical indication for each scan was not available.

### 3.1. Classification Task

We evaluated classification performance on the held-out evaluation dataset using both our ResNet50 and Vision Transformer (ViT) architectures. All reported metrics utilize a decision threshold of 0.50 (unless otherwise stated). 95% confidence intervals have also been calculated to account for statistical uncertainty.

As can be seen in Table 3, ResNet50 achieved strong overall performance, with an accuracy of 98.5% (95% CI: [96.7, 100.0]), a precision of 97.4% (95% CI: [94.2, 100.0]), a recall of 100% (95% CI: [100.0, 100.0]; the zero-width interval reflects the absence of false negatives across all 2,000 bootstrap resamples, consistent with the confusion matrix), and an F1-score of 98.7% (95% CI: [97.0, 100.0]), together with an AUROC of 0.999 ± 0.0002 and an AUPRC of 0.999 ± 0.0001. The confusion matrix for the ResNet50 classifier (Table 4) shows a low number of false positives and no observed false negatives.

The ViT model achieved slightly lower but still competitive performance, with an accuracy of 96.6% (95% CI: [93.8, 99.0]), a precision of 99.1% (95% CI: [97.1, 100.0]), a recall of 94.8% (95% CI: [90.4, 98.3]), and an F1-score of 96.9% (95% CI: [94.3, 99.0]), with an AUROC of 0.996 ± 0.0032 and an AUPRC of 0.997 ± 0.0021. While ViT maintained exceptional performance, its confusion matrix (Table 5) shows a higher false-negative rate relative to ResNet50, suggesting that convolutional architectures may be better suited for NCCT-based stroke classification in limited-data and low-contrast conditions.

A McNemar test comparing the two models yielded a p-value of 0.34, indicating that the performance difference was not statistically significant.

To confirm our models’ robustness in early and subtle stroke cases, we analyzed their performance in cases with lesion volumes of less than 5 mL. In this scenario, ResNet50 in particular achieved perfect detection performance with no false negatives. Accuracy, precision, recall, and F1-score all remained at 100% for these cases, despite the low contrast and limited spatial extent of the infarcts. These smaller lesions make up the majority of our stroke-positive cases, which further supports the robustness of our models in clinically challenging domains.

Our ResNet50 model demonstrated excellent probabilistic calibration as well, with an expected calibration error (ECE) of 0.06, and a Brier score of 0.02. Meanwhile, ViT exhibited a good Brier score of 0.09, but it was substantially more poorly calibrated and notably under-confident, with predicted probabilities concentrated at intermediate values rather than near 0 or 1 (ECE = 0.25). This behavior seems to be consistent with the existing knowledge that transformers have trouble making accurate and confident predictions in limited data settings. For further context, we have also provided calibration curves for both models in the Supplementary Material (Figures S.1 and S.2).

Across a variety of thresholds ranging from 0.35 to 0.75, ResNet50 performed more robustly than ViT, with high recall and stable overall performance at all thresholds. In contrast, ViT’s performance degraded more noticeably as the decision threshold changed, indicating that ViT is more sensitive to threshold selection. It is worth noting that both models performed remarkably well at moderate thresholds between 0.45 and 0.65. These findings suggest that ResNet50 is likely better suited for deployment in clinical settings where higher confidence thresholds may be preferred. Detailed quantitative results for the full threshold sweep are provided in the Supplementary Material (Tables S.1 and S.2).

### 3.2. Segmentation Task

While we saw extremely strong performance for our classification models on our small-lesion dataset, segmentation proved to be considerably more difficult. This is an expected result, since ischemic lesions on NCCT often exhibit very low contrast, and the majority of cases in our dataset consist of small-volume lesions, making precise segmentation challenging.

We evaluated segmentation performance using four architectures as described earlier: a classic attention U-Net, a modified attention U-Net (AUIS), a 3D U-Net, and a dynamic U-Net.

Throughout all architectures, segmentation accuracy depended heavily on lesion volume. Small lesions consistently resulted in lower Dice scores regardless of model choice. Meanwhile, larger lesions were segmented with substantially higher accuracy.

Figure 5 plots the Dice score vs lesion size in the test set for our classic Attention U-Net model. Most lesions smaller than 5 mL (aside from a small number of outliers) resulted in near-zero Dice scores, indicating that the model was largely unable to meaningfully segment infarcts in this size range. As lesion volume increased, segmentation performance improved substantially, with Dice scores rising steadily up to approximately 60 mL. Beyond this range, performance became more variable, most likely due to the scarcity of very large lesions in the dataset. Overall, this figure clearly indicates that NCCT-based segmentation depends heavily on lesion size, though it should be noted that near-zero Dice scores on small lesions may also partly reflect label noise introduced during MRI coregistration, where registration errors of even 1-2 mm can result in misaligned ground truth masks for very small lesions. Similar figures for all three other models are provided in the Supplementary Material (Figures S.3–S.5).

**Figure 5.**
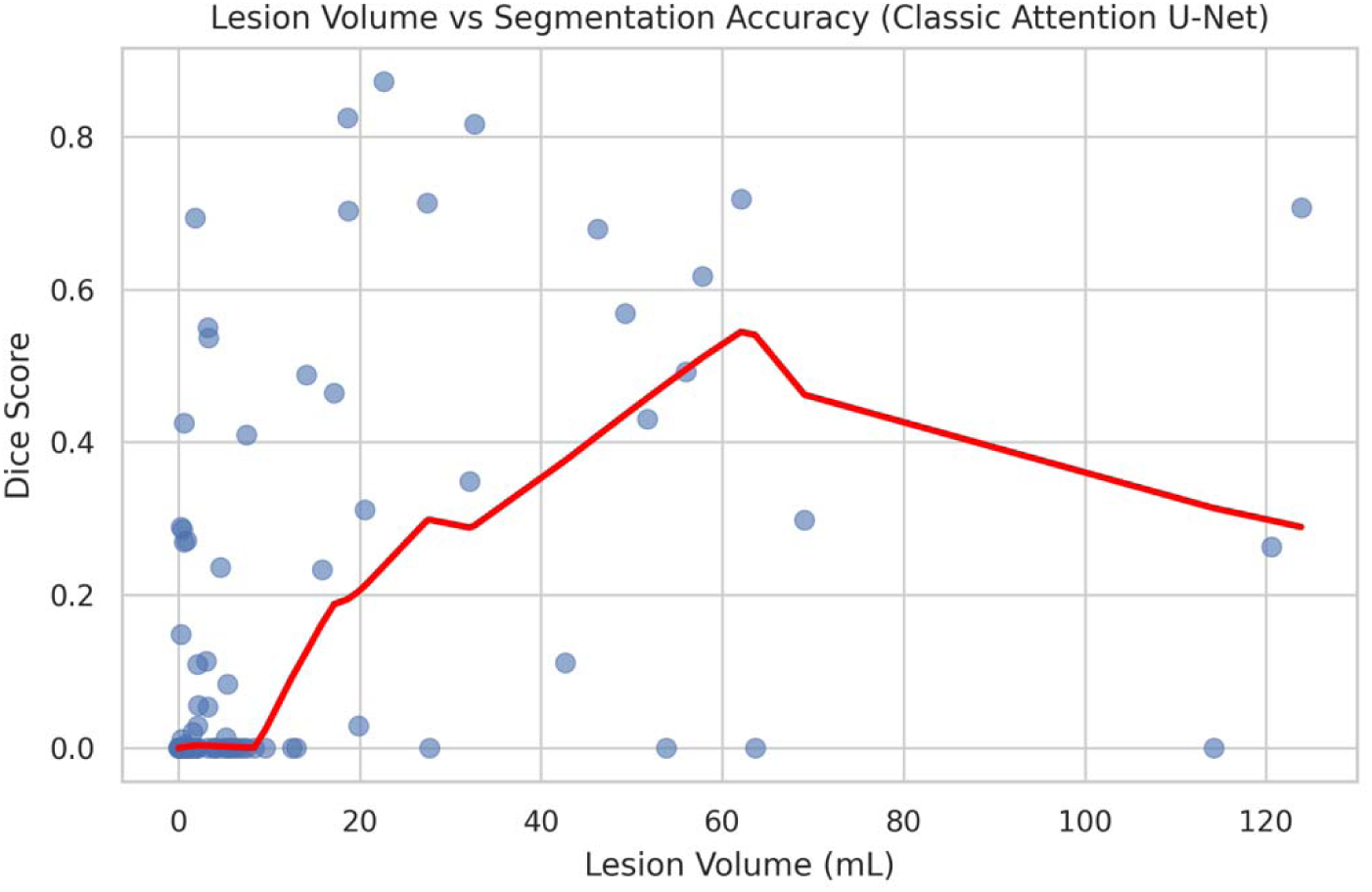
Relationship between lesion volume (mL = milliliters) and Dice coefficient across the evaluation set for the classic Attention U-Net. Larger lesions show higher segmentation accuracy.

Despite these limitations, we also observed cases in which the models achieved high-quality segmentations for larger lesions, demonstrating the feasibility of segmentation under favorable conditions. Our classic Attention U-Net model obtained the highest single subject Dice score of 0.87, while AUIS (0.82), dynamic U-Net (0.80), and 3D U-Net (0.77) returned comparable peak performances. While these peak Dice scores do not reflect the average performance of our models on our dataset, they demonstrate that segmentation is feasible for high-volume lesions. Figure 6 showcases two examples of predictions from our AUIS model for larger lesions, further indicating that the primary challenge is the prevalence of smaller lesions in our dataset rather than model capability.

**Figure 6.**
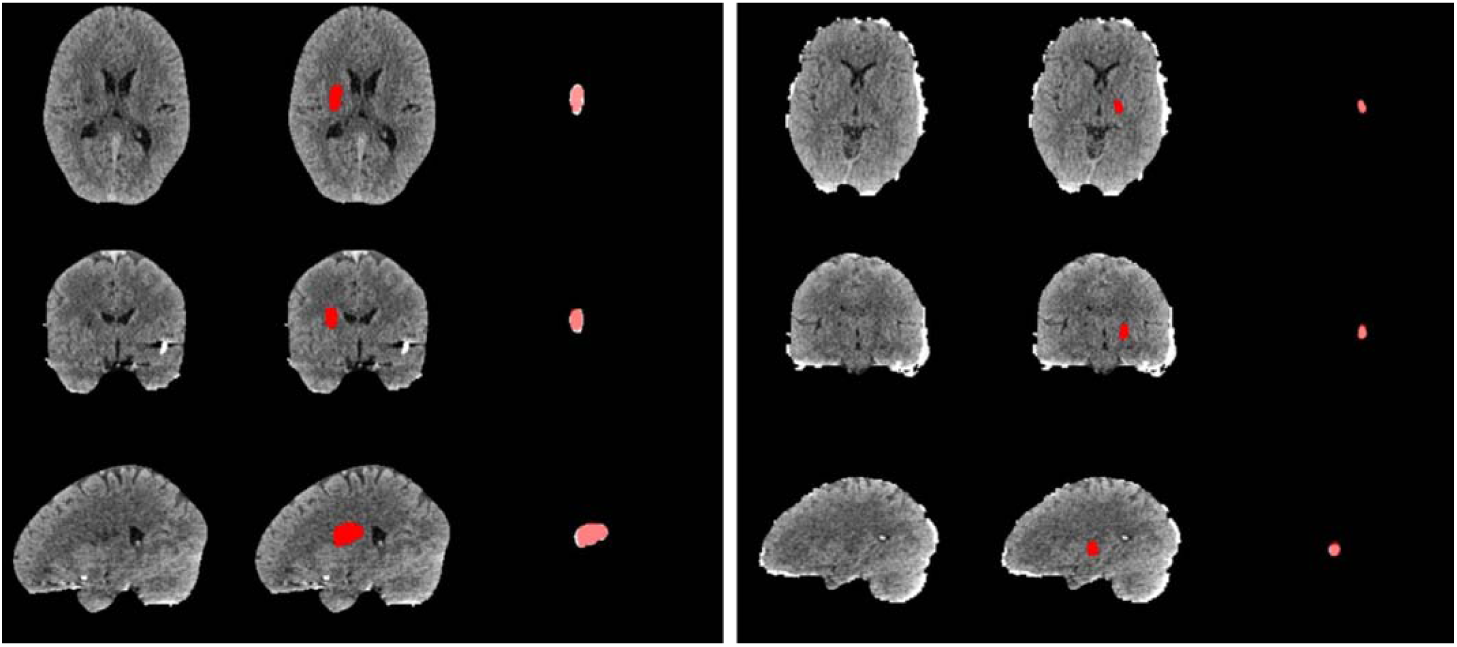
Example segmentation predictions from the AUIS model for two larger lesions. First column: original NCCT scan. Second column: true lesion overlaid in red. Third column: true lesion (red) and model prediction (white) overlaid together.

For completeness, detailed quantitative metrics including mean Dice scores evaluated across multiple lesion volume thresholds are reported in the Supplementary Material (Tables S.3–S.6).

## 4. Discussion

We find that deep learning models are capable of accurate early AIS classification on NCCT data, and that this extends to the detection of small infarcts (<5 mL). We utilize ResNet50 and Vision Transformer (ViT) architectures to extract features and classify the scans in an effort to provide an accurate and timely stroke detection system. We provide a systematic comparison of various deep learning architectures for lesion segmentation on NCCT, demonstrating that lesion size is a primary limiting factor for accurate segmentation. We characterize the strengths of NCCT-based classification and the current limitations of NCCT-based segmentation for future methodological and data-driven improvement.

Deep learning aligns well with neuroimaging tasks because it can learn hierarchical, nonlinear spatial-temporal features directly from high-dimensional brain images, capturing subtle distributed patterns that classical hand-engineered methods often miss. In the realm of AIS detection, prior research using deep learning has focused on MRI-based methods using fluid-attenuated inversion recovery (FLAIR) [18, 19] and DWI [9, 20, 21, 22]. Similarly, prior work has explored deep learning methods applied to CTA/CTP [23, 24, 25] imaging, whose constraints have been discussed. With regards to stroke classification on NCCT, Kuang et al. [26] proposed a hybrid CNN-Transformer architecture with circular feature interaction and bilateral difference learning for NCCT lesion segmentation, outperforming 17 state-of-the-art methods and achieving Dice scores of 61.4% and 46.7% on two AIS datasets. More recently, Heo et al. [27] developed and externally validated a modified 3D U-Net trained on 2,214 multicenter patients, demonstrating 75.3% sensitivity and 79.1% specificity for lesion detection on NCCT, though notably, lesions smaller than 0.5 mL were excluded from evaluation due to NCCT’s inherent resolution limitations.

### 4.1. Classification Task

Our classification results demonstrate that automated lesion detection using a volume-level classification approach, in which slice-level features are extracted and aggregated across the full scan, is a viable approach to expedite the stroke treatment pipeline. Both our models were able to detect lesions on NCCT scans with high accuracy. ResNet50 demonstrated strong probabilistic calibration with an ECE of 0.06 and a Brier score of 0.02. ViT, while competitive in accuracy, showed substantially poorer calibration (under-confident, with predictions concentrated at intermediate probabilities) with an ECE of 0.25, consistent with the known data requirements of transformer architectures in limited-data settings.

Importantly, our models performed robustly on lesions smaller than 5 mL. This confirms that these models are not only viable on highly curated, large-lesion NCCT scans but also on smaller and subtler lesions that would be encountered in the very early stages of the stroke pathogenesis. This is clinically significant, since small, early lesions are the cases where manual NCCT interpretation would be most difficult and where missed detection would be the most costly.

Recent large-scale work has explicitly excluded very small lesions from evaluation due to NCCT resolution constraints [27], making robust performance in this regime a largely unaddressed clinical gap that our results begin to address.

The apparent tension between strong classification performance and near-zero segmentation Dice on sub-5 mL lesions should be discussed. These results, which may appear contradictory at first, rather reflect the fundamentally different information requirements of the two tasks.

Segmentation demands precise voxel-level boundaries, requiring sufficient local contrast and spatial extent to reconstruct an accurate mask. Classification requires only a global signal of abnormality across the entire volume. Subtle ischemic changes that are insufficient for reliable voxel-level boundaries may nonetheless produce detectable shifts in global image statistics, including subtle asymmetries or density changes, that a volumetric classifier can learn to exploit.

Our threshold sensitivity analysis indicates that our models are able to maintain their performance in a wide range of thresholds. This result is crucial because it allows clinicians and hospitals to tune the model to their preference. A model with a higher decision threshold would allow for fewer false positives to occur, thus reducing the potential costs of further treatment when it is unnecessary. On the other hand, a model with a lower decision threshold would allow for fewer false negatives and ensure that any patient with a possible stroke is treated effectively. Operating points may vary by clinician or institution, thus our models’ robustness across thresholds suggests potential utility across different clinical operating points, though prospective validation in diverse clinical settings would be necessary before deployment. We note that these results were obtained on a held-out evaluation set from a single institution, and external validation across multiple sites would be necessary to confirm generalizability. Visual explainability analysis such as Grad-CAM was not performed and represents an important direction for future work. Additionally, while all scans were acquired at a single institution under a consistent clinical protocol, scanner-level matching between stroke-positive and stroke-negative cases could not be confirmed, and systematic acquisition differences between cohorts represent a potential confounding factor that future work should address through explicit cohort matching.

While a decision threshold of 0.5 was chosen for primary reporting to maintain consistency with other literature in the field, it is essential to recognize that in a clinical setting, different thresholds might be considered more suitable. In cases where an automated classification pipeline is treated as a signal for downstream treatment, a higher decision threshold might be preferred in order to avoid false positives, which could lead to wasted resources and clinician attention. In contrast, a lower decision threshold might also be favorable in order to avoid missed detections. While the ultimate decision of which threshold to utilize should be made by the clinicians themselves, it is crucial that our models are well calibrated regardless of the chosen threshold. Therefore, we conducted a threshold sensitivity analysis to evaluate our models’ stability across a range of decision thresholds.

### 4.2. Segmentation Task

Our segmentation results reveal fundamental limitations of lesion segmentation on NCCT. Across all four tested architectures, the Dice score tracked closely with lesion volume. Small lesions overwhelmingly produced low Dice scores, regardless of the chosen model. As lesion size increased, we saw a substantial improvement in segmentation quality.

The prevalence of small lesions that strengthened our classification results was unfortunately the primary weakness encountered by our segmentation models. Even the classic Attention U-Net and AUIS, which showed the most promise in segmenting large lesions, were unable to meaningfully segment the majority of small lesions in our dataset. This finding suggests that lesion size and the inherent contrast limitations of NCCT are significant contributors to segmentation difficulty, alongside potential confounding factors such as label and registration noise introduced during the MRI coregistration process used to generate ground truth masks.

This interpretation is supported by recent work, as Kuang et al. [26], using a sophisticated hybrid CNN-Transformer architecture, achieved Dice scores of 46.7% on a private clinical dataset with a more favorable size distribution, which is comparable to our models’ performance on lesions above 10 mL. The authors also directly identify performance on smaller lesions as a limitation of their models, suggesting that this pattern holds across model architectures.

Small lesions on NCCT often have extremely low contrast and poorly defined boundaries. In many cases, there is not enough visual information in the scan to accurately segment the lesion. Unlike classification, segmentation demands more precise spatial boundaries. This explains why our classification models succeeded on the same cases that challenged our segmentation models.

### 4.3. Takeaways

These results reveal a clear divide between the current viability of classification and segmentation tasks on NCCT. Classification of strokes on NCCT is accurate and clinically useful, even with the small-lesion infarcts that dominated our dataset. Segmentation, while possible on larger lesions with good accuracy, is more sensitive to lesion size and struggles with earlier, smaller strokes.

While a larger dataset with more diverse lesion sizes may improve segmentation performance, our results suggest that the barrier of lesion size is difficult to surpass when trying to precisely segment lesions on NCCT. Future progress could utilize various strategies, such as aiming for general localization of the stroke rather than precise segmentation, incorporating a larger dataset with more large lesions for initial training, and developing more powerful model architectures.

Until then, NCCT-based classification may offer the most immediate clinical value while segmentation remains an open problem. A model that reliably informs the clinician of stroke presence in its earliest stages, even without precise localization, represents a meaningful advancement in the automated stroke diagnosis pipeline.

## 5. Conclusion

We investigated the feasibility of deep learning-based AIS classification and segmentation using a volume-level approach on NCCT scans, specifically on a large dataset dominated by small, early-stage lesions. Our results show a divergence between the two tasks on NCCT.

Classification of lesions is reliable, with our models able to accurately detect AIS even when lesions are small and subtle. The models maintain their performance across a variety of decision thresholds and for lesion volumes below 5 mL, suggesting that NCCT-based classification is viable and clinically valuable for stroke diagnosis.

Meanwhile, segmentation remains a substantially more difficult problem. Across all architectures, performance strongly depended on lesion size. Small lesions were rarely correctly segmented, while larger lesions were segmented with higher accuracy. This indicates that the primary limitation for stroke segmentation lies in the inherent low contrast and ambiguity of small lesions on NCCT.

Future work may benefit from reframing segmentation objectives towards localization rather than precise segmentation, incorporating larger and more diverse datasets, and integrating more powerful attention-based models. However, in the near term, the ability to reliably detect acute ischemic stroke on NCCT in its earliest stages represents a clinically meaningful step forward, particularly in settings where advanced imaging is unavailable or time constraints are severe.

## Supporting information

All Tables

## Data Availability

The dataset used in this study contains protected patient data and is not publicly available due to patient privacy constraints. The dataset was compiled from clinical scans at Johns Hopkins University under IRB approval number IRB00164427. The study was conducted in accordance with the Declaration of Helsinki. Informed consent was waived by the institutional review board due to the retrospective nature of the study and the use of de-identified data in compliance with HIPAA regulations. The complete source code for the models is publicly available on GitHub under the following link: https://github.com/AnshGoyal1123/Acute-Stroke-Detection

## CRediT Author Contributions

**Ansh Goyal:** Conceptualization, Data curation, Formal analysis, Investigation, Methodology, Software, Visualization, Writing – original draft, Writing – review and editing.

**Robert David Stevens:** Conceptualization, Funding acquisition, Project administration, Methodology, Resources, Supervision, Writing – review and editing

## Acknowledgements

The authors wish to thank Brenda Johnson, Assistant Director of the Johns Hopkins Hospital Comprehensive Stroke Center, as well as Feng-Chiao Lee, Rongxi Yi, Xinyuan Fang, Ike Zhang, and Yanlin Wu from the Laboratory of Computational Intensive Care Medicine at Johns Hopkins University for their contributions to the early stages of this project.

## References

1. J.A. Chalela, C.S. Kidwell, L.M. Nentwich, M. Luby, J.A. Butman, A.M. Demchuk, M.D. Hill, N. Patronas, L. Latour, S. Warach, Magnetic resonance imaging and computed tomography in emergency assessment of patients with suspected acute stroke: a prospective comparison, Lancet 369 (9558) (2007) 293–298.

2. W.J. Powers, A.A. Rabinstein, T. Ackerson, O.M. Adeoye, N.C. Bambakidis, K. Becker, J. Biller, M. Brown, B.M. Demaerschalk, B. Hoh, E.C. Jauch, C.S. Kidwell, T.M. Leslie-Mazwi, B. Ovbiagele, P.A. Scott, K.N. Sheth, A.M. Southerland, D.V. Summers, D.L. Tirschwell, Guidelines for the early management of patients with acute ischemic stroke: 2019 update to the 2018 guidelines for the early management of acute ischemic stroke: a guideline for healthcare professionals from the American Heart Association/American Stroke Association, Stroke 50 (12) (2019) e344–e418.

3. C.D. Kulathilake, J. Udupihille, S.P. Abeysundara, A. Senoo, Deep learning–driven multi-class classification of brain strokes using computed tomography, Eur. J. Radiol. 187 (2025) 112109.

4. J. Sun, G.L. Ju, Y.H. Qu, H.H. Xie, H.X. Sun, S.Y. Han, Y.F. Li, X.Q. Jia, Q. Yang, Deep learning for segmenting ischemic stroke infarction in non-contrast CT scans by utilizing asymmetry, Clin. Neuroradiol. (2025) (epub ahead of print).

5. A. Tuladhar, S. Schimert, D. Rajashekar, H. Kniep, J. Fiehler, N. Forkert, Automatic segmentation of stroke lesions in non-contrast computed tomography datasets with convolutional neural networks, IEEE Dataport, 2020.

6. T. Fuchigami, S. Akahori, T. Okatani, Y. Li, A hyperacute stroke segmentation method using 3D U-Net integrated with physicians’ knowledge for NCCT, in: Proc. SPIE Medical Imaging 2020: Computer-Aided Diagnosis, vol. 11314, SPIE, 2020, p. 113140G.

7. A. Fedorov, R. Beichel, J. Kalpathy-Cramer, J. Finet, J.-C. Fillion-Robin, S. Pujol, C. Bauer, D. Jennings, F. Fennessy, M. Sonka, J. Buatti, S. Aylward, J.V. Miller, S. Pieper, R. Kikinis, 3D Slicer as an image computing platform for the quantitative imaging network, Magn. Reson. Imaging 30 (9) (2012) 1323–1341.

8. A.C. Evans, A.L. Janke, D.L. Collins, S. Baillet, Brain templates and atlases, NeuroImage 62 (2) (2012) 911–922.

9. C.-F. Liu, J. Hsu, X. Xu, S. Ramachandran, V. Wang, M. Miller, A. Hillis, A. Faria, M. Wintermark, S. Warach, G. Albers, S. Davis, J. Grotta, W. Hacke, D.-W. Kang, C. Kidwell, W. Koroshetz, K. Lees, M. Lev, M. Luby, Deep learning-based detection and segmentation of diffusion abnormalities in acute ischemic stroke, Commun. Med. 1 (2021) 61.

10. S. Klein, M. Staring, K. Murphy, M.A. Viergever, J.P.W. Pluim, Elastix: a toolbox for intensity-based medical image registration, IEEE Trans. Med. Imaging 29 (1) (2010) 196–205.

11. K. He, X. Zhang, S. Ren, J. Sun, Deep residual learning for image recognition, in: Proc. IEEE Conf. Comput. Vis. Pattern Recognit. (CVPR), 2016, pp. 770–778.

12. A. Dosovitskiy, L. Beyer, A. Kolesnikov, D. Weissenborn, X. Zhai, T. Unterthiner, M. Dehghani, M. Minderer, G. Heigold, S. Gelly, J. Uszkoreit, N. Houlsby, An image is worth 16×16 words: transformers for image recognition at scale, arXiv:2010.11929, 2020.

13. O. Oktay, J. Schlemper, L.L. Folgoc, M. Lee, M. Heinrich, K. Misawa, K. Mori, S. McDonagh, N.Y. Hammerla, B. Kainz, B. Glocker, D. Rueckert, Attention U-Net: learning where to look for the pancreas, arXiv:1804.03999, 2018.

14. Ö. Çiçek, A. Abdulkadir, S.S. Lienkamp, T. Brox, O. Ronneberger, 3D U-Net: learning dense volumetric segmentation from sparse annotation, in: Proc. MICCAI 2016, in: Lecture Notes in Computer Science, vol. 9901, Springer, 2016, pp. 424–432.

15. F. Isensee, P.F. Jaeger, S.A.A. Kohl, J. Petersen, K.H. Maier-Hein, nnU-Net: a self-configuring method for deep learning-based biomedical image segmentation, Nat. Methods 18 (2) (2021) 203–211.

16. M.J. Cardoso, W. Li, R. Brown, N. Ma, E. Kerfoot, Y. Wang, B. Murrey, A. Myronenko, C. Zhao, D. Yang, V. Nath, Y. He, Z. Xu, A. Hatamizadeh, W. Zhu, Y. Liu, M. Zheng, Y. Tang, I. Yang, A. Feng, MONAI: an open-source framework for deep learning in healthcare, arXiv:2211.02701, 2022.

17. H. Iqbal, PlotNeuralNet v1.0.0, Zenodo, 2018.

18. M.T. Duong, J.D. Rudie, J. Wang, L. Xie, S. Mohan, J.C. Gee, A.M. Rauschecker, Convolutional neural network for automated FLAIR lesion segmentation on clinical brain MR imaging, Am. J. Neuroradiol. 40 (8) (2019) 1282–1290.

19. S. Khezrpour, H. Seyedarabi, S.N. Razavi, M. Farhoudi, Automatic segmentation of the brain stroke lesions from MR FLAIR scans using improved U-Net framework, Biomed. Signal Process. Control 78 (2022) 103978.

20. L. Chen, P. Bentley, D. Rueckert, Fully automatic acute ischemic lesion segmentation in DWI using convolutional neural networks, NeuroImage Clin. 15 (2017) 633–643.

21. R. Zhang, L. Zhao, W. Lou, J.M. Abrigo, V.C.T. Mok, W.C. Chu, D. Wang, L. Shi, Automatic segmentation of acute ischemic stroke from DWI using 3-D fully convolutional DenseNets, IEEE Trans. Med. Imaging 37 (9) (2018) 2149–2160.

22. K. Ito, H. Kim, S.-L. Liew, A comparison of automated lesion segmentation approaches for chronic stroke T1-weighted MRI data, Hum. Brain Mapp. 40 (16) (2019) 4669–4685.

23. J. Bertels, D. Robben, D. Vandermeulen, P. Suetens, A. Crimi, S. Bakas, H. Kuijf, F. Keyvan, M. Reyes, T. Van Walsum, Contra-lateral information CNN for core lesion segmentation based on native CTP in acute stroke, in: Proc. MICCAI 2019, in: Lecture Notes in Computer Science, vol. 11383, Springer, 2019, pp. 263–270.

24. G. Wang, T. Song, Q. Dong, M. Cui, N. Huang, S. Zhang, Automatic ischemic stroke lesion segmentation from computed tomography perfusion images by image synthesis and attention-based deep neural networks, Med. Image Anal. 65 (2020) 101787.

25. M. Soltanpour, R. Greiner, P. Boulanger, B. Buck, Improvement of automatic ischemic stroke lesion segmentation in CT perfusion maps using a learned deep neural network, Comput. Biol. Med. 137 (2021) 104849.

26. H. Kuang, Y. Wang, J. Liu, J. Wang, Q. Cao, B. Hu, W. Qiu, J. Wang, Hybrid CNN-Transformer network with circular feature interaction for acute ischemic stroke lesion segmentation on non-contrast CT scans, IEEE Trans. Med. Imaging 43 (6) (2024) 2303–2316.

27. J.N. Heo, W.-S. Ryu, J.-W. Chung, C.K. Kim, J.-T. Kim, M. Lee, D. Kim, L. Sunwoo, J.M. Ospel, N. Singh, H.-J. Bae, B.J. Kim, Automated ischemic stroke lesion detection on non-contrast brain CT: a large-scale clinical feasibility test, Front. Neurosci. 19 (2025) 1643479.

