## Supplementary material for "Acute Ischemic Stroke Detection on Non-Contrast CT: A Deep Learning Approach": All Tables

Ansh Goyal, Robert David Stevens

**Contents**

**Tables	2**

Table 1. Clinical Demographic Information	2

Table 2. Lesion Volume Distribution	3

Table 3. Classification Performance on Evaluation Set	3

Table 4. Confusion Matrix for ResNet50	4

Table 5. Confusion Matrix for ViT	5

### Tables

| **Variable** | **Stroke(N=578)** | **Non-Stroke(N=463)** |
| --- | --- | --- |
| **Age, mean (SD), years** | 63.3 (13.2) | 60.3 (5.9) |
| **Male, %** | 53.3% | 56.9% |
| **First NIHSS, mean (SD)** | 4.8 (5.4) | N/A |
| **Time, stroke onset to NCCT**  **h, mean (SD)** | 6.6 (9.5) | N/A |
| **Time, stroke onset to NCCT**  **h, median (IQR)** | 3.5 (2.4 – 5.1) | N/A |
| **Time, stroke onset to MRI**  **h, mean (SD)** | 20.0 (15.3) | N/A |
| **Time, stroke onset to MRI**  **h, median (IQR)** | 16.3 (13.3 – 20.1) | N/A |
| **In-hospital death, %** | 1.4% | 0% |
| **mRS at 3 months, mean (SD)** | 1.91 (1.76) | N/A |

#### Table 1.

Sample characteristics for the stroke-positive and stroke-negative groups. Onset-to-imaging intervals calculated from registry timestamps; cases lacking complete date/time precision were imputed using the median value of cases with precise timestamps. Mean and median both reported given right-skewed underlying distribution. Mean mRS on 456 of 578 patients for whom data were available. NIHSS, NIH stroke scale; NCCT, non-contrast CT; mRS, modified Rankin Scale

| **Lesion Volume Range (mL)** | **Number of Scans** | **Percentage of Scans** |
| --- | --- | --- |
| **0 – 1** | 213 | 36.8% |
| **1.1 – 5** | 162 | 28.0% |
| **5.1 – 10** | 49 | 8.5% |
| **10.1 – 30** | 85 | 14.7% |
| **30.1 – 60** | 37 | 6.4% |
| **>60** | 32 | 5.5% |
| **Total** | **578** | **100%** |

#### Table 2.

Lesion volume distribution across all 578 stroke-positive cases in the dataset. The majority of cases (64.9%) fall below 5 mL, reflecting the early-stage and subtle ischemic presentations that characterize this dataset.

| **Model Name** | **Accuracy** | **Recall** | **Precision** | **AUROC** | **AUPRC** |
| --- | --- | --- | --- | --- | --- |
| **ResNet50 [19]** | 98.5 ± 0.85 | 100.0 ± 0.00 | 97.4 ± 1.49 | 0.999 ± 0.0002 | 0.999 ± 0.0001 |
| **ViT [20]** | 96.6 ± 1.24 | 94.8 ± 2.07 | 99.1 ± 0.90 | 0.996 ± 0.0032 | 0.997 ± 0.0021 |

#### Table 3.

Classification performance for both classification models on the evaluation set (n = 209)

|  | **Predicted Stroke** | **Predicted Non-Stroke** |
| --- | --- | --- |
| **True Stroke** | 116 | 0 |
| **True Non-Stroke** | 3 | 90 |

#### Table 4.

Confusion matrix for ResNet50 on the Evaluation Set (Threshold = 0.50, n = 209)

|  | **Predicted Stroke** | **Predicted Non-Stroke** |
| --- | --- | --- |
| **True Stroke** | 110 | 6 |
| **True Non-Stroke** | 1 | 92 |

#### Table 5.

Confusion matrix for ViT on the Evaluation Set (Threshold = 0.50, n = 209)
