## Supplementary Materials for "Acute Ischemic Stroke Detection on Non-Contrast CT: A Deep Learning Approach"

Ansh Goyal, Robert David Stevens

**Supplementary Material**

**Contents**

**Classification Threshold Sweep Data	2**

Table S.1. Threshold Sensitivity Analysis for ResNet50	2

Table S.2. Threshold Sensitivity Analysis for ViT	3

**Classification Model Calibration Plots	4**

Figure S.1. Calibration plot for ResNet50	4

Figure S.2. Calibration plot for ViT	5

**Segmentation Lesion Size vs. Dice Score Plots	6**

Figure S.3. Lesion size vs. Dice score for AUIS	6

Figure S.4. Lesion size vs. Dice score for 3D U-Net	7

Figure S.5. Lesion size vs. Dice score for Dynamic U-Net	7

**Segmentation Performance Across Volume Thresholds	8**

Table S.3. Segmentation Performance of AUIS	8

Table S.4. Segmentation Performance of Classic Attention U-Net	8

Table S.5. Segmentation Performance of 3D U-Net	9

Table S.6. Segmentation Performance of Dynamic U-Net	9

### Classification Threshold Sweep Data

Below is the threshold sensitivity analysis for the ResNet50 model. The confusion matrices for all thresholds past 0.55 look identical, which tells us that the model is quite confident in most of its positive predictions. This is a good sign, especially considering that the model performs exceptionally well on our dataset.

| **Threshold** | **Acc** | **Prec** | **Rec** | **F1** | **TP** | **TN** | **FP** | **FN** |
| --- | --- | --- | --- | --- | --- | --- | --- | --- |
| 0.35 | 0.943 | 0.906 | 1.000 | 0.951 | 116 | 81 | 12 | 0 |
| 0.45 | 0.971 | 0.951 | 1.000 | 0.975 | 116 | 87 | 6 | 0 |
| 0.55 | 0.986 | 0.975 | 1.000 | 0.987 | 116 | 90 | 3 | 0 |
| 0.65 | 0.986 | 0.975 | 1.000 | 0.987 | 116 | 90 | 3 | 0 |
| 0.75 | 0.986 | 0.975 | 1.000 | 0.987 | 116 | 90 | 3 | 0 |

#### Table S.1. Threshold Sensitivity Analysis for ResNet50 on the Evaluation Set

Below is the same threshold sensitivity analysis for the ViT model. This model seems to deteriorate as we raise the decision threshold, indicating that it is less confident in its predictions. It is commonly known that transformer models require larger amounts of data than other models to attain similar performance, but it is unclear if that is what is causing this difference in confidence.

| **Threshold** | **Acc** | **Prec** | **Rec** | **F1** | **TP** | **TN** | **FP** | **FN** |
| --- | --- | --- | --- | --- | --- | --- | --- | --- |
| 0.35 | 0.914 | 0.871 | 0.991 | 0.927 | 115 | 76 | 17 | 1 |
| 0.45 | 0.962 | 0.950 | 0.983 | 0.966 | 114 | 87 | 6 | 2 |
| 0.55 | 0.952 | 1.000 | 0.914 | 0.955 | 106 | 93 | 0 | 10 |
| 0.65 | 0.866 | 1.000 | 0.759 | 0.863 | 88 | 93 | 0 | 28 |
| 0.75 | 0.651 | 1.000 | 0.371 | 0.541 | 43 | 93 | 0 | 73 |

#### Table S.2. Threshold Sensitivity Analysis for Vision Transformer (ViT) on the Evaluation Set

### Classification Model Calibration Plots

Below is the calibration plot for the ResNet50 model. The sharp bimodal nature of the plot indicates the model's confidence in its predictions is extremely high.

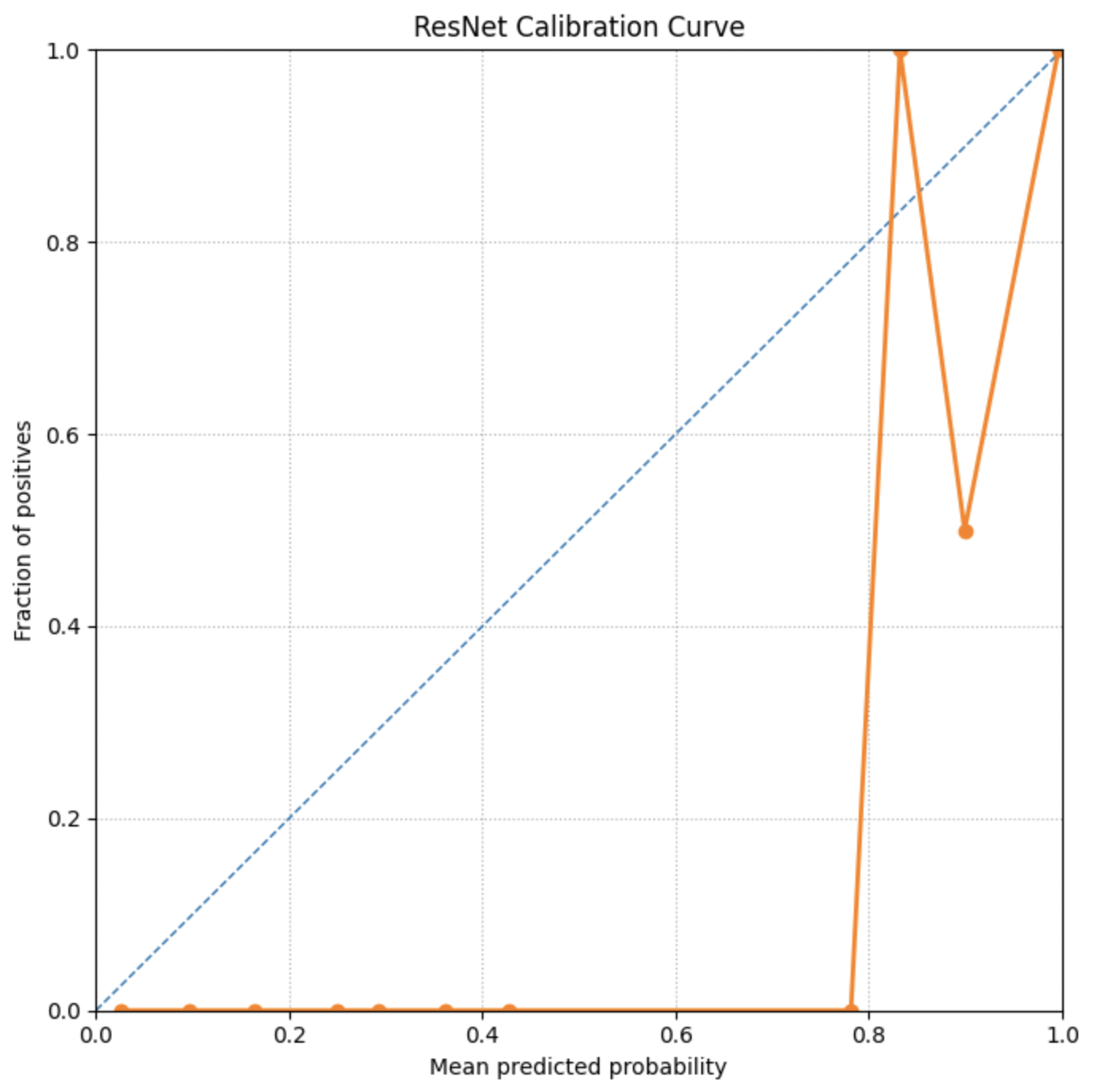

#### Figure S.1. Calibration plot for ResNet50.

Below is the calibration plot for the ViT model. This plot indicates that unlike the ResNet50 model, ViT is less confident in its predictions, which is supported by the threshold analysis shown above.

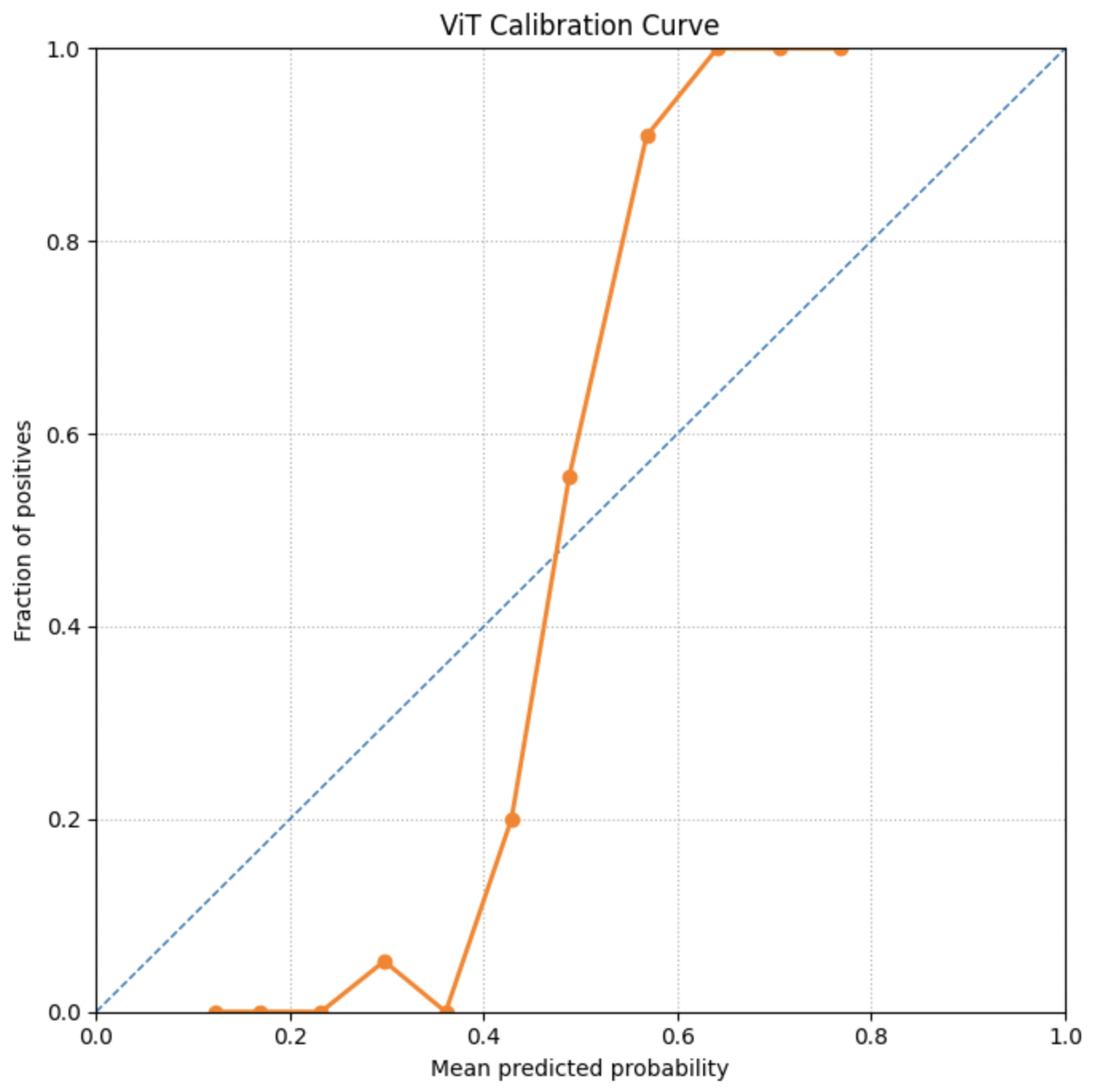

#### Figure S.2. Calibration plot for ViT.

### Segmentation Lesion Size vs. Dice Score Plots

Below are the lesion size vs. Dice score plots for the other three models tested for segmentation.

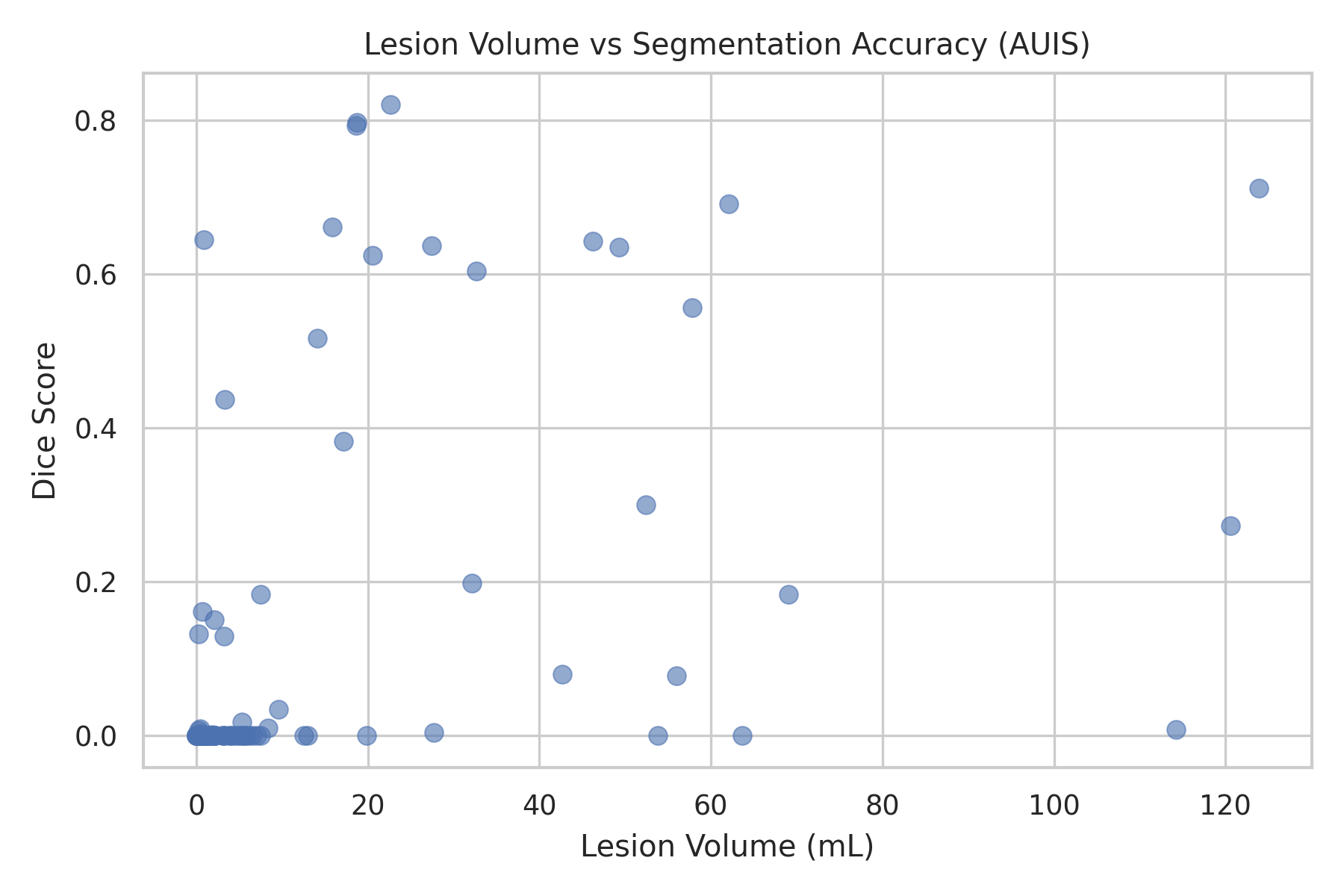

#### Figure S.3. Lesion size vs. Dice score plotted for AUIS.

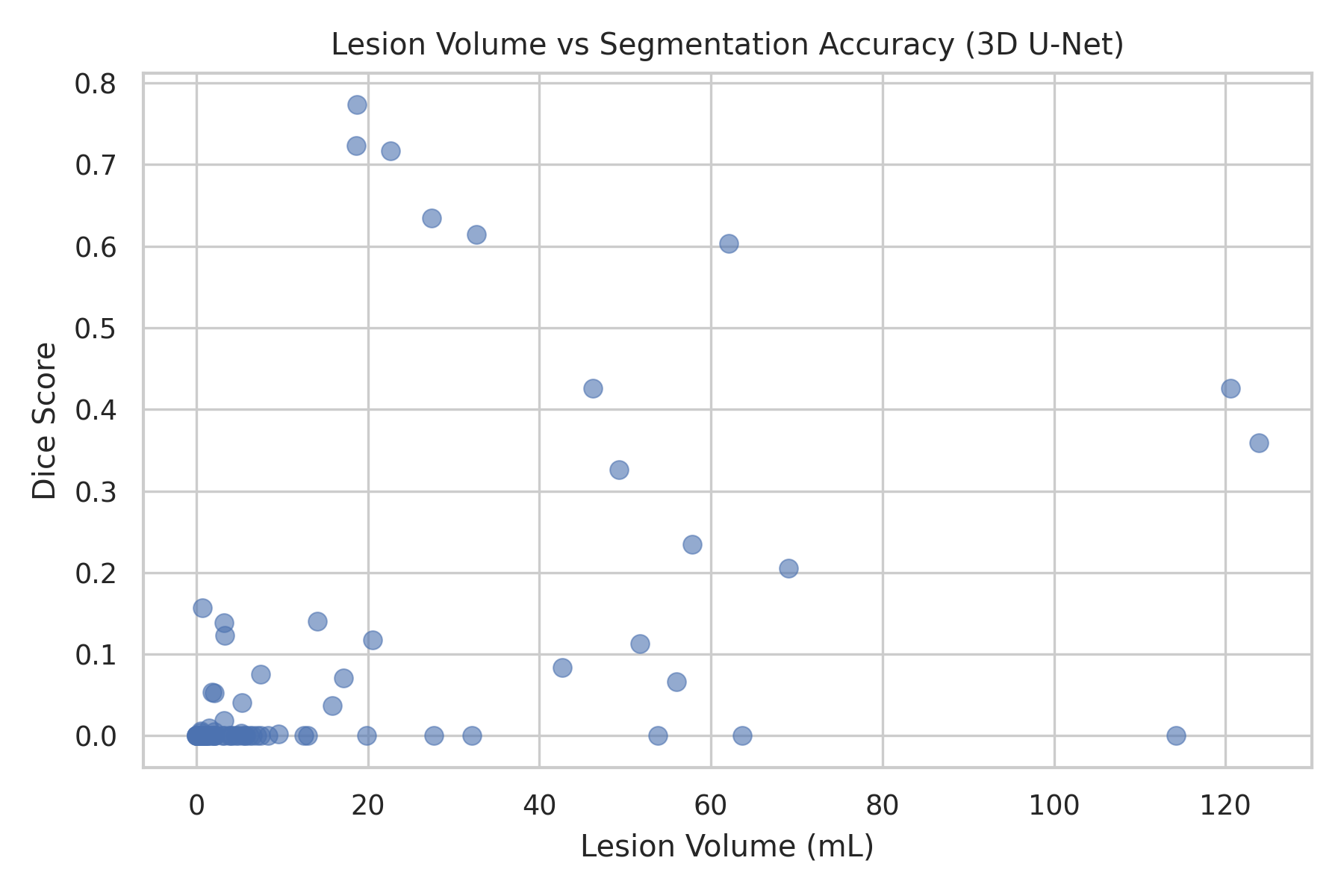

#### Figure S.4. Lesion size vs. Dice score plotted for 3D U-Net.

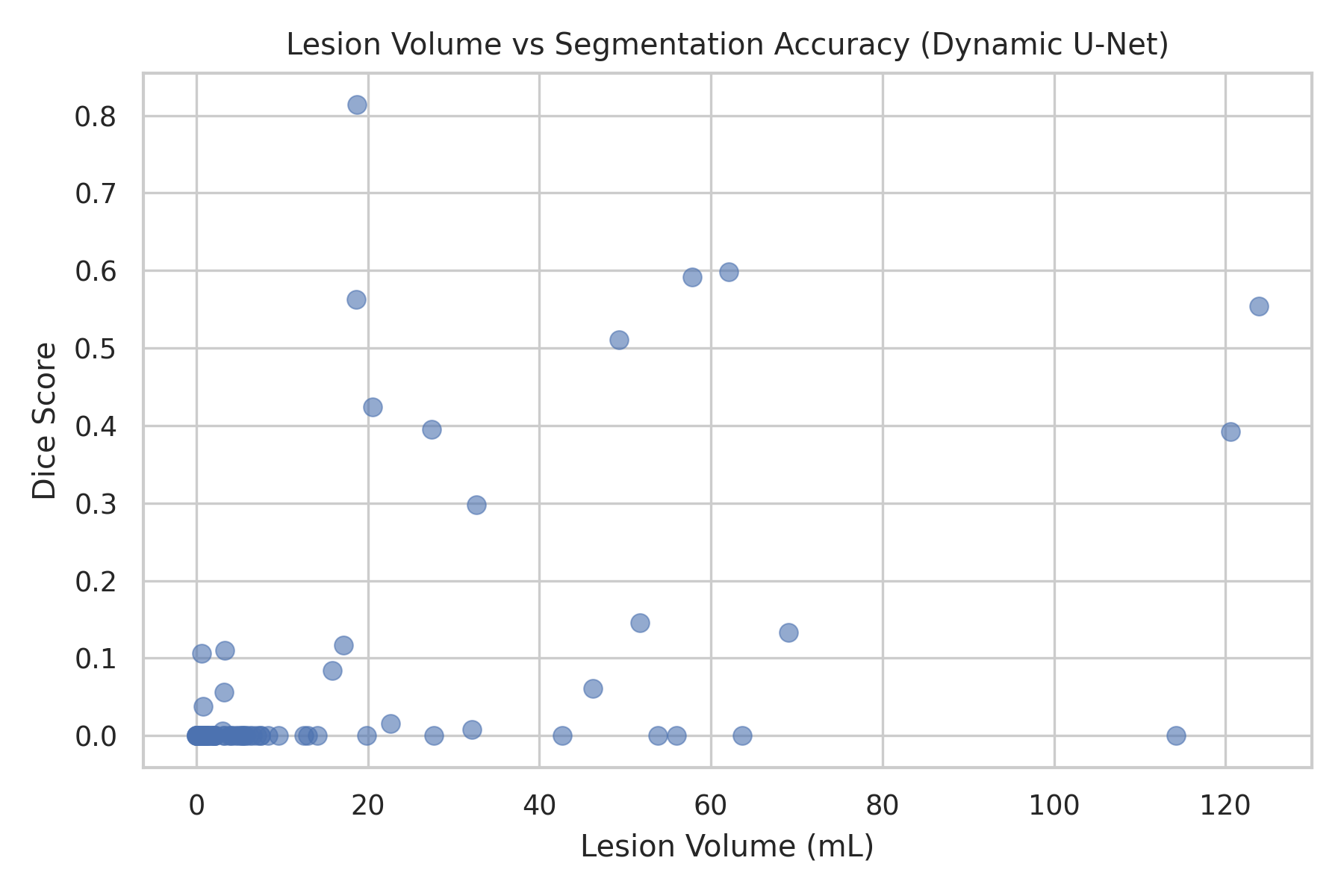

#### Figure S.5. Lesion size vs. Dice score plotted for Dynamic U-Net.

### Segmentation Performance Across Volume Thresholds

The tables below showcase the mean and standard deviations of Dice scores for each model for all test cases with lesion volume above the provided threshold.

| **Threshold (mL)** | **Mean Dice** | **Dice Std** |
| --- | --- | --- |
| 0 | 0.104 | 0.223 |
| 5 | 0.268 | 0.302 |
| 10 | 0.378 | 0.302 |
| 30 | 0.331 | 0.269 |
| 60 | 0.311 | 0.292 |

#### Table S.3. Segmentation Performance of AUIS Across Testing Volume Thresholds

| **Threshold (mL)** | **Mean Dice** | **Dice Std** |
| --- | --- | --- |
| 0 | 0.132 | 0.236 |
| 5 | 0.287 | 0.303 |
| 10 | 0.396 | 0.297 |
| 30 | 0.403 | 0.274 |
| 60 | 0.331 | 0.293 |

#### Table S.4. Segmentation Performance of Classic Attention U-Net Across Testing Volume Thresholds

| **Threshold (mL)** | **Mean Dice** | **Dice Std** |
| --- | --- | --- |
| 0 | 0.063 | 0.165 |
| 5 | 0.174 | 0.246 |
| 10 | 0.247 | 0.265 |
| 30 | 0.231 | 0.210 |
| 60 | 0.266 | 0.221 |

#### Table S.5. Segmentation Performance of 3D U-Net Across Testing Volume Thresholds

| **Threshold (mL)** | **Mean Dice** | **Dice Std** |
| --- | --- | --- |
| 0 | 0.052 | 0.150 |
| 5 | 0.146 | 0.230 |
| 10 | 0.211 | 0.250 |
| 30 | 0.220 | 0.236 |
| 60 | 0.280 | 0.247 |

#### Table S.6. Segmentation Performance of Dynamic U-Net Across Testing Volume Thresholds
